# Exploring healthcare experiences and access needs in unplanned hospital admissions for Inflammatory Bowel Disease: A multi-perspective qualitative study

**DOI:** 10.64898/2026.05.26.26353596

**Authors:** Rachel L Hawkins, Charlotte Cotterill, Samantha McCormick, Ian Kellar, Alan J Lobo, Fiona Sampson

## Abstract

**Background:** Unplanned hospital admissions in Inflammatory Bowel Diseases (IBD) account for nearly three-quarters of IBD inpatient stays in the United Kingdom. Although costly to services and distressing for patients, research exploring experiences and potential drivers of admissions is limited. We undertook a qualitative study to explore the healthcare experiences and access needs of people with IBD who had unplanned admissions, along with their caregivers and clinicians.

**Methods:** Semi-structured interviews with 25 participants from a single tertiary IBD service in England (*17* people with IBD, *3* informal caregivers, *5* clinicians) were conducted. We applied thematic framework analysis, guided by the Candidacy Framework, and worked with *2* patient and public contributors to generate final themes.

**Results:** We identified four themes: ‘Difficulties in *Identifying flares and asserting severity before admission’*, summarised the prevailing uncertainty in identifying a flare and access to timely IBD care. *‘Navigating a disjointed healthcare system’*, highlighted how lack of care plans and systemic barriers can delay access. *‘Emergency care access challenges’* highlighted the gaps in emergency and inpatient care during flares. Whilst *‘fighting for care and individual advocacy needs’*, described the persistent assertion for care that may disproportionally impact access to vulnerable groups, also highlighting the importance of positive interpersonal relationships.

**Conclusions:** Individual, interpersonal and healthcare factors across the patient pathway were perceived to shape access to care in unplanned IBD admissions. Potentially reducing admissions requires proactive strategies, including the integration of patient education, monitoring tools, establishment of specialist rapid-access pathways, and formal psychological support to address barriers to access.

**Summary:** Research exploring experiences of IBD admissions is limited. We conducted qualitative interviews to explore patient, caregiver and clinician perspectives revealing individual, interpersonal and healthcare factors that shaped access and experiences of care before and after admissions.

**What is already known?:** Unplanned IBD admissions are frequent, costly, and distressing for patients. Whilst recognised as markers of suboptimal service quality, research into the healthcare experiences and systemic drivers behind these admissions remains limited.

**What is new here?:** This is the first study to explore healthcare access leading up to unplanned IBD admissions from multiple perspectives. We show that poor knowledge of flares and services, patient and clinicians’ uncertainty during initial contact, and the overall impermeability of healthcare can delay access and contribute to unplanned IBD admissions. Furthermore, persistent assertion for care is required in IBD that may disproportionally impact access for vulnerable groups.

**How can this study help patient care?:** Findings from this study can directly inform health service interventions to reduce IBD admissions including improved patient flare and navigation information, patient monitoring tools and establishing specialist rapid-access clinics to provide alternatives to A&E. Furthermore, embedding formal psychological support can address the emotional barriers to accessing care.

## Introduction

Inflammatory Bowel Diseases (IBD) are chronic and unpredictable conditions including Crohn’s Disease (CD) and Ulcerative Colitis (UC) that affect over 500,000 people in the United Kingdom (UK) (1). The relapsing and remitting nature of IBD requires access to long-term specialist IBD care (2). However, widespread workforce shortages and a decline in the quality of care imposes barriers to this access (1,3). Unplanned hospital admissions are a frequent and costly outcome in IBD management (1,4,5) with a profound impact on psychological wellbeing (6,7).

Unplanned IBD admissions potentially driven by inequalities in IBD care (8,9) are recognised as a marker of suboptimal service quality, often representing the culmination of missed opportunities for timely intervention in the community (10). However, current health inequalities research in IBD is limited, with further research needed exploring healthcare experiences of people with IBD across ethnicity, gender and sexual identity and disability (8). Whilst improved patient-centred care is a key feature of current IBD management guidelines (2,11), inclusive and diverse evidence to design services remains underdeveloped. Understanding the patient experience is therefore an important step for service improvement (12) and reducing admissions.

Focused exploratory research in the context of IBD hospital admissions is also limited (6) and healthcare interventions to reduce admissions may misalign with the needs of people with IBD. Embedding clinician and caregiver perspectives can also support comparison of views, potentially adding new perspectives to healthcare improvement (13).

We undertook a qualitative study to explore the healthcare experiences and access needs of diverse people with IBD who had unplanned hospital admissions for their IBD, also interviewing their informal caregivers and IBD clinicians (consultants, nurses, surgeons) leading their care to build a broader picture of future health service improvement. The Candidacy Framework (14), which guides understanding of how vulnerable populations access healthcare services, supported the organisation and interpretation of the data (15).

## Method

The Consolidated Criteria for Reporting Qualitative Research (16) is used for the reporting of this study.

### Design

A qualitative design using semi-structured interviews explored the perspectives of people with IBD, caregivers and IBD clinicians. Qualitative interviews supported the unfolding of diverse experiences of IBD healthcare related to unplanned admissions in IBD (17). A topic guide (appendix D) was developed using Candidacy (14), to explore how the journey of accessing IBD healthcare leading up to, during and after an unplanned IBD admission. The Candidacy Framework (14) is a widely used framework for understanding how vulnerable populations access healthcare services. It has been applied across different health contexts for understanding healthcare access, including cancer (18), rheumatoid arthritis (19), and emergency care (20). Candidacy has also been applied in understanding inequities in IBD care related to unplanned admissions (21). Table 1 maps the constructs of candidacy to IBD care (supplementary file A).

**Table 1.** Stages of Candidacy adapted for inflammatory bowel disease healthcare.

| **Candidacy Construct** | **General description from Dixon-Woods et al. (2006)** | **Adapted description to IBD healthcare** |
| --- | --- | --- |
| **Identification of Candidacy** | How people come to recognise symptoms as needing medical intervention. | PLwIBD identifies themselves as a candidate for IBD care. This is influenced by their social contexts, prior experience and knowledge of bowel symptoms, IBD and healthcare. |
| **Navigation of IBD services** | To access services, people must be aware of them and have available practical resources to access them. | Requires awareness of care options, mobilisation of resources (e.g. transport to clinics) and understanding of how the service manages IBD. For example, knowing that IBD flare advice lines are a first point of call during a flare. |
| **Permeability of IBD services** | Refers to the ease at which people can use health services, which is impacted by the organisation of services. This may include accessing appointments or referrals. | Represents PLwIBD journey through the system. The permeability of IBD services, i.e. the availability of and barriers to expert advice in primary or secondary care. |
| **Appearance at services** | When appearing at services people must have the competence to articulate their issue and assert their claim for medical intervention. | PLwIBD must have the skills and confidence to present themselves to clinicians. This requires competencies in articulating IBD needs, and an understanding of IBD treatment options. |
| **Adjudications by healthcare professionals** | Professional judgments are made by the healthcare professional about candidacy and their eligibility for care. This then strongly influences subsequent access to | Candidacy also requires judgement and decision making of IBD health professionals. The clinician identifies the patient as a candidate for interventions. Requires assessment of disease |
|  | healthcare and interventions. | status and severity of IBD flares. |
| **Offers of, and resistance to services** | Some people may choose to refuse offers or there may be resistance to healthcare and interventions such as referrals and medications. | This stage involves decision-making of PLwIBD to accept new treatments (e.g. biologics, steroids, surgery) and attend appointments. This is influenced by individual, social and organisational factors including trust in health care professionals and institutions. |
| **Operating conditions and local production of candidacy** | These include factors at the societal and macro levels that influence candidacy. For example, the availability of local resources and the relational aspects between the healthcare provider and the patient. | Represents the impact prior and current relationships PLwIBD have with their IBD team which impact Candidacy. Access to IBD care is also impacted by the availability of specialist IBD resources, and local and national policies that impact funding of services. |
| *PLwIBD = People living with inflammatory bowel disease* |  |  |

### Participants and recruitment

This study was set in a regional South Yorkshire adult IBD service that provides tertiary IBD care to approximately 5000 patients through dedicated IBD outpatient clinics with gastroenterologists and IBD specialist nurses, and an IBD-nurse led advice line. Two population groups were targeted for recruitment:

1. Adults (aged >16 years) with a diagnosis of IBD and a recorded unplanned IBD hospital admission from Feb 2022 to March 2025. All participants were provided with the choice to bring an informal caregiver to the interview.
2. The IBD specialist leading the care at the time of the unplanned IBD admission (e.g. IBD consultant, nurse, surgeon). Informed consent was obtained from patient participants to contact the named professional at the end of the interview.

A REC approved study flyer signed letter from an IBD consultant [AJL], and a participant information sheet (PIS) approved by the Patient Oversight Committee (POC) prior to ethical approval (patient and public involvement detail is attached to supplementary file B) was distributed via email, handed to patients during clinics and posted via mail. Four waves of targeted recruitment were undertaken with people with IBD, listed below:

1. Identified and invited via an existing database, made available via the AWARE-IBD patient cohort (22), who had consented to be contacted for future research opportunities.
2. IBD clinicians shared the recruitment materials with patients deemed eligible using the sampling frame during clinic appointments. A sampling frame was developed (See supplementary C), following a review of existing patient experience measure (PREM) data available via AWARE-IBD (22).
3. In-person recruitment on the inpatient ward. One female researcher (RH) liaised with gastroenterologists who distributed the PIS to IBD patients admitted as an unplanned IBD admission. Following consent, RH visited the patient on the ward to share information and was invited to contact the researcher after hospital discharge to take part.
4. Targeted recruitment of diverse ethnic groups admitted through the service’s IBD advice line. Thirteen participants were contacted via email or post to take part in the study.

IBD clinicians were recruited via a consultant signed invitation email. All patient participants responded to an optional consent question to contact their healthcare professional involved in the unplanned IBD admission for interview. All patient participants consented. Fourteen clinicians were invited to take part in the study.

Informed consent, either verbal or written, was obtained from all participants prior to taking part in the study. Upon consent, all participants were allocated a study ID number and pseudonym.

### Data collection

In-person and remote semi-structured interviews were conducted between August 2024 - March 2025 by one white British female doctoral researcher (RH) with prior qualitative research experience. Two separate, but structurally similar topics guides were developed and structured using the Candidacy Framework (Supplementary D) and reviewed by the POC.

Interviews with people with IBD and informal caregivers were conducted in person in a private university room (n=4) and remote via telephone or GoogleMeet (n=13). All clinician interviews were conducted remotely. All interviews were conducted in English. Three people with IBD attended the interview with an informal caregiver. During interviews, a timeline was established about their experience leading up to, during and after the unplanned admission. The topic guide was developed to enable flexibility, and remained open to discussing other important aspects of care.

All interviews were audio-recorded and transcribed verbatim by RH removing all person and identifiable information from the data. During interviews, the lead researcher (RH) made reflexive notes which were maintained throughout data collection.

### Data analysis

A whole group analysis was conducted, treating the data produced by the participant groups as a whole (23). Data analysis was led by RH, with the support of CC, FS and two public contributors (SMc, RS). RH and CC collaborated using a shared NVivo file and Miro board.

We used a hybrid deductive and inductive thematic framework analysis approach to data analysis, using NVivo for data organisation (17). First, all transcripts were read line-by-line by RH for data familiarisation. In addition, FS read two full transcripts and CC read three full transcripts, which were coded and discussed in meetings. The framework approach (15) was applied, using predetermined organising codes developed from the Candidacy Framework (deductive approach) in which codes related to the constructs were organised in NVivo. Inductive codes were also coded outside of the framework (inductive approach) (15,24).

Following coding of all interviews, framework matrices were used to visualise and compare perspectives across the dataset cases as a subgroup analysis (23). Analysis of the framework matrices supported the development of themes across different cases (patient/caregiver/clinician). These typologies are useful by showing how particular views or experiences may attach to particular groups and can help inform recommendations for improving IBD patient care (23). Initial themes were agreed following discussions at team meetings (RH, FS, IK, CC), and were finalised at the final PPI analysis meeting.

Embedding PPI during the analysis boosted reciprocal learning opportunities and improved the quality, rigor and process of reflexivity in the data analysis through triangulating with multiple perspectives (25). The full detail of the studies approach to embedding PPI in the analysis is outlined in supplementary file B.

### Ethical approval

The study received ethical approval from the Health and Care Research Support Centre, Wales Research Ethics Committee 3 (REC reference 21/WA/0264).

## Results

### Participants

We interviewed twenty-five participants (17 people with IBD, 3 informal caregivers, 4 consultant gastroenterologists and 1 IBD specialist nurse). Table 2 presents their demographics. Participants with IBD were admitted via the IBD advice line, self-attendance at accident and emergency (A&E), calls to emergency services and during GP appointments. To preserve anonymity, ethnicity data has been broadened and the relationships of caregivers are not reported.

**Table 2.** Participant demographic information.

| Characteristic | Participant number (%) |  |  |
| --- | --- | --- | --- |
|  | People with IBD (*n*=17) | Informal *Caregivers* (*n*=3) | IBD clinicians (*n*=5) |
| *Gender* |  |  |  |
| Male | 8 (47) | 1 (33.4) | 1 (20) |
| Female | 8 (47) | 2 (66.6) | 4 (80) |
| Non-binary | 1 (6) | - | - |
| *Professional* |  |  |  |
| Gastroenterologist | - | - | 4 (80) |
| IBD Specialist Nurse | - | - | 1 (20) |
| *Age years (mean)* | 46.5 years | - | - |
| *IBD subtype* |  | - |  |
| Crohn’s Disease | 9 (52.9) |  |  |
| Ulcerative Colitis | 8 (47.1) |  |  |
| *Index of Multiple Deprivation (IMD)* |  |  |  |
| Decile 1 & 2 (most deprived) | 5 (29.4) |  |  |
| Decile 3 & 4 | 3 (17.6) |  |  |
| Decile 5 & 6 | 6 (35.2) |  |  |
| Decile 7 & 8 | 1 (5.9) |  |  |
| Decile 9 & 10 (least deprived) | 2 (11.8) |  |  |
| ***Ethnicity*** |  |  |  |
| White British or Irish | 12 (70.5) |  |  |
| Other White | 1 (5.9) |  |  |
| Mixed Ethnicity | 2 (11.8) |  |  |
| Pakistani | 1 (5.9) |  |  |
| Indian | 1 (5.9) |  |  |
| ***Physical Disability ("Yes")*** | 4 (23.5) |  |  |
| ***Learning disability ("Yes")*** | 1 (5.9) |  |  |
| ***Neurodiversity ("Yes")*** | 2 (11.8) |  |  |
| **Comorbid physical/mental health diagnosis** |  |  |  |
| 1 physical/mental health condition | 5 (29.4) |  |  |
| > 1 physical/mental health conditions | 4 (23.5) |  |  |
| ***Route of unplanned IBD admission*** |  |  |  |
| Via the IBD advice line (direct to hospital ward or A&E) | 9 (52.9) |  |  |
| Self attendance to A&E | 4 (23.5) |  |  |
| Call to emergency services | 2 (11.8) |  |  |
| GP admission to same day emergency care | 1 (5.9) |  |  |
| Admitted during a hospital procedure | 1 (5.9) |  |  |
| Where "-" is entered, data was not collected |  |  |  |

We identified four themes with seven subthemes which represent people with IBD healthcare access and experiences. The four themes represent the interconnectedness of the patient journey through healthcare up until hospital admission, highlighting that navigating care doesn’t necessarily occur in discrete steps. Figure 1 maps these themes through use of a patient journey timeline which illustrates barriers to healthcare access impacting unplanned IBD admissions, using the Candidacy Framework (14). Table 3 presents additional data (Supplementary file E).

**Table 3.**
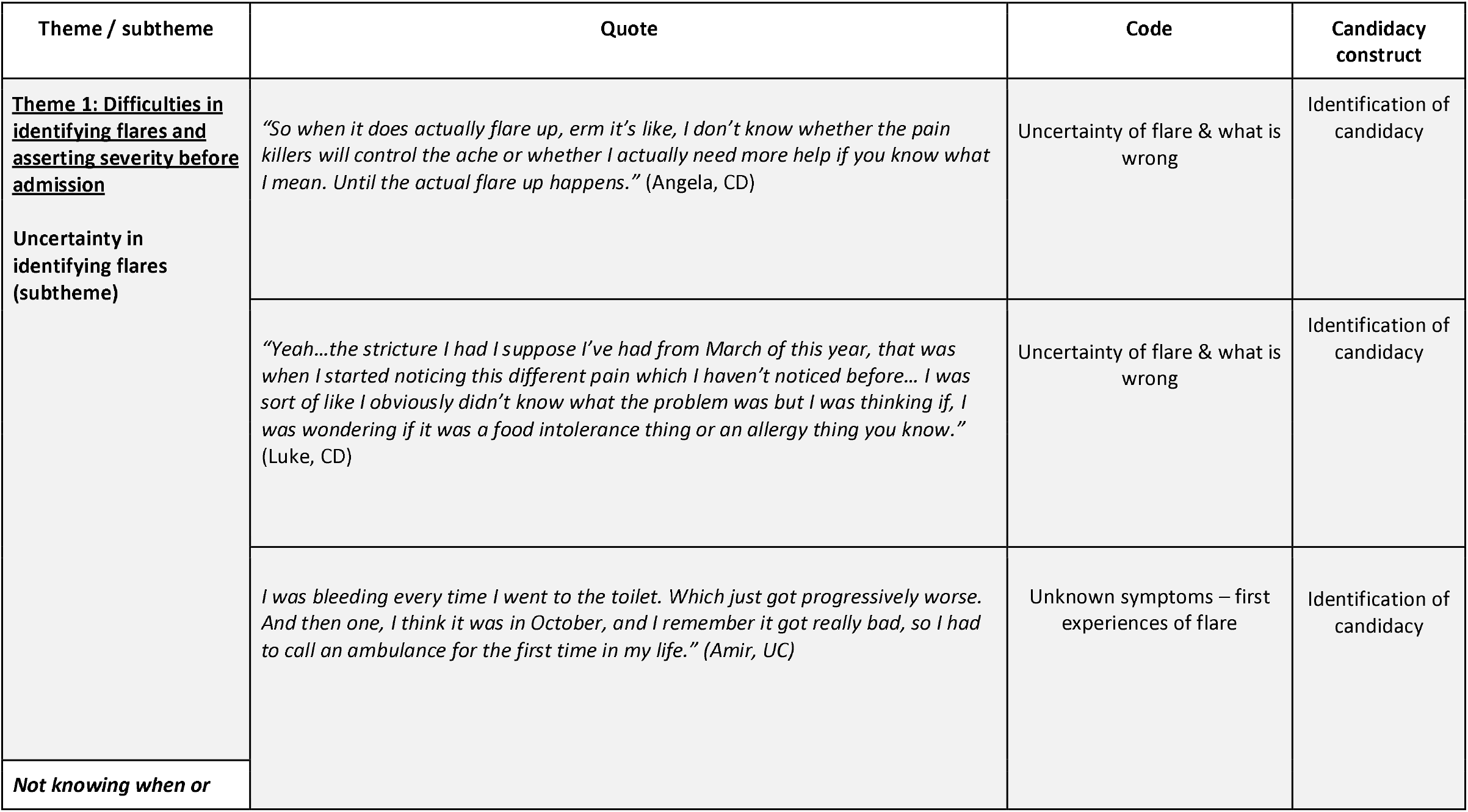
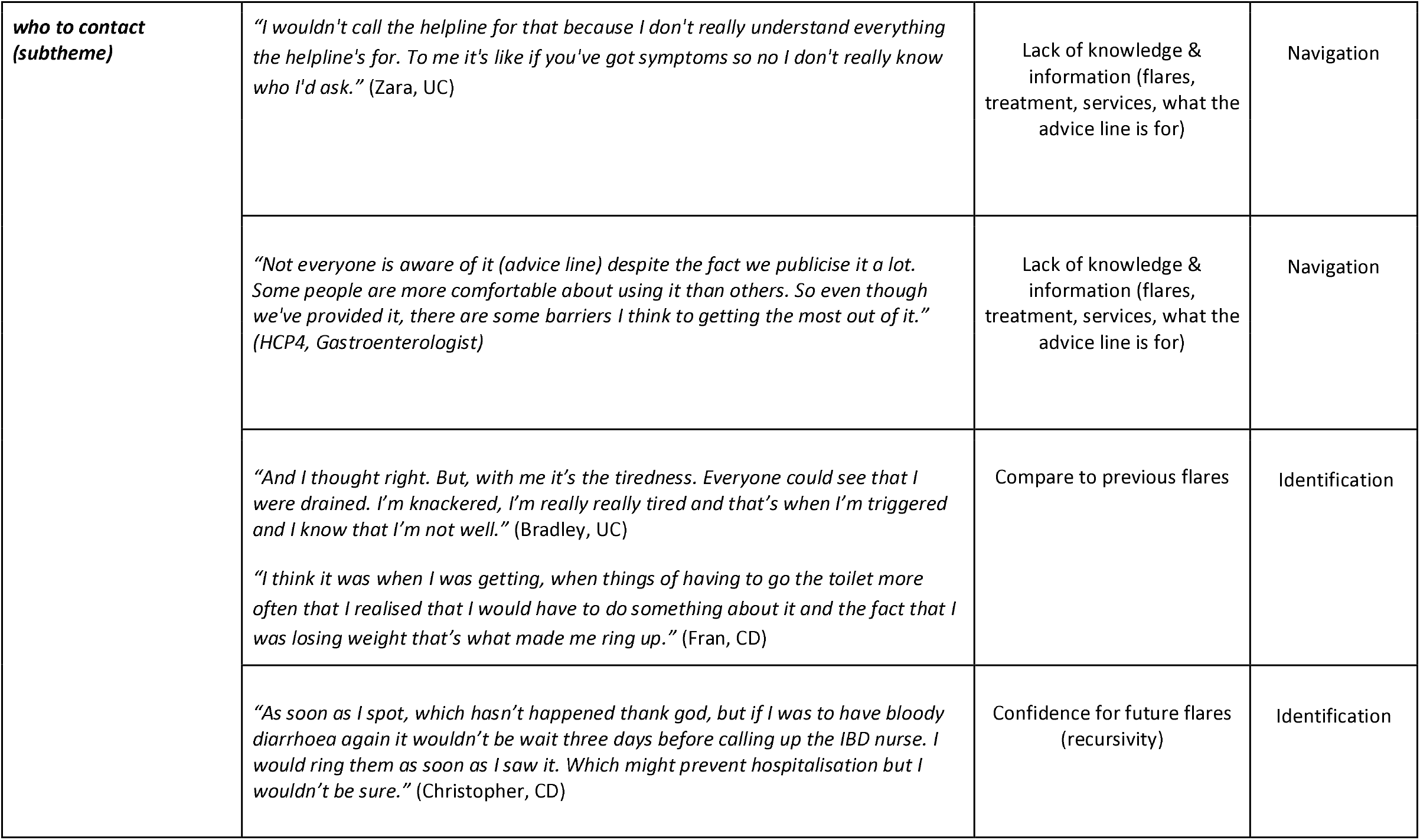
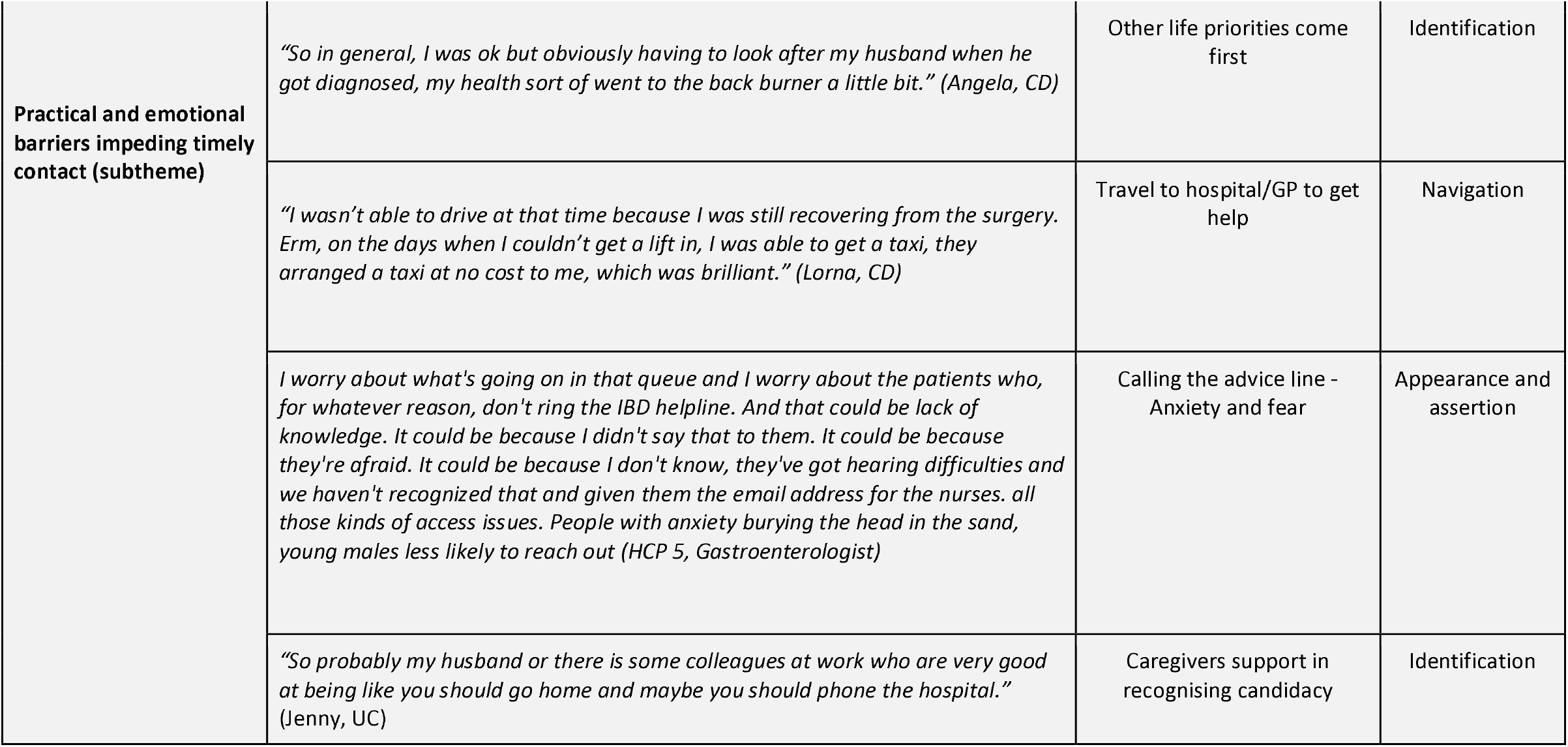
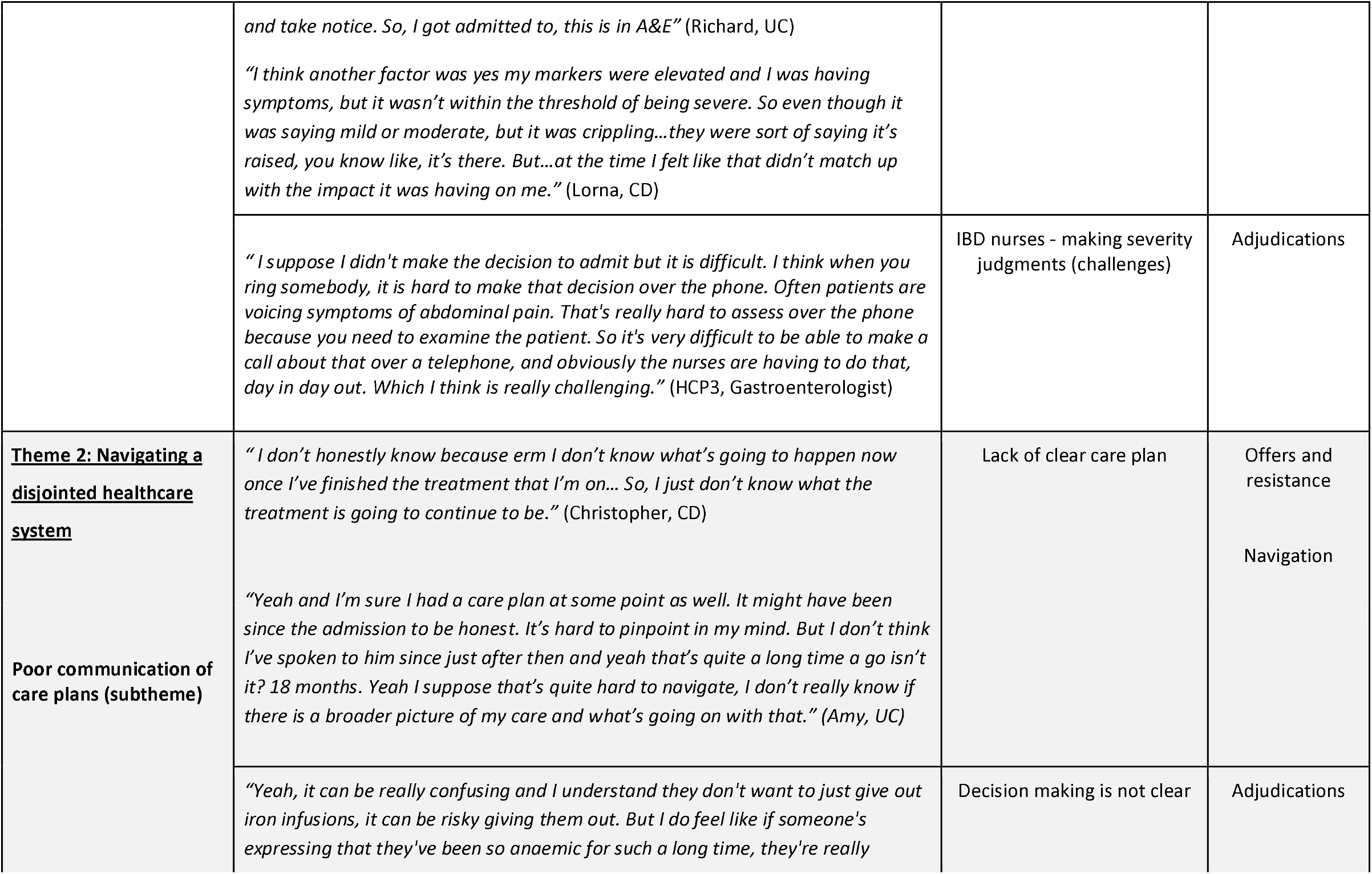
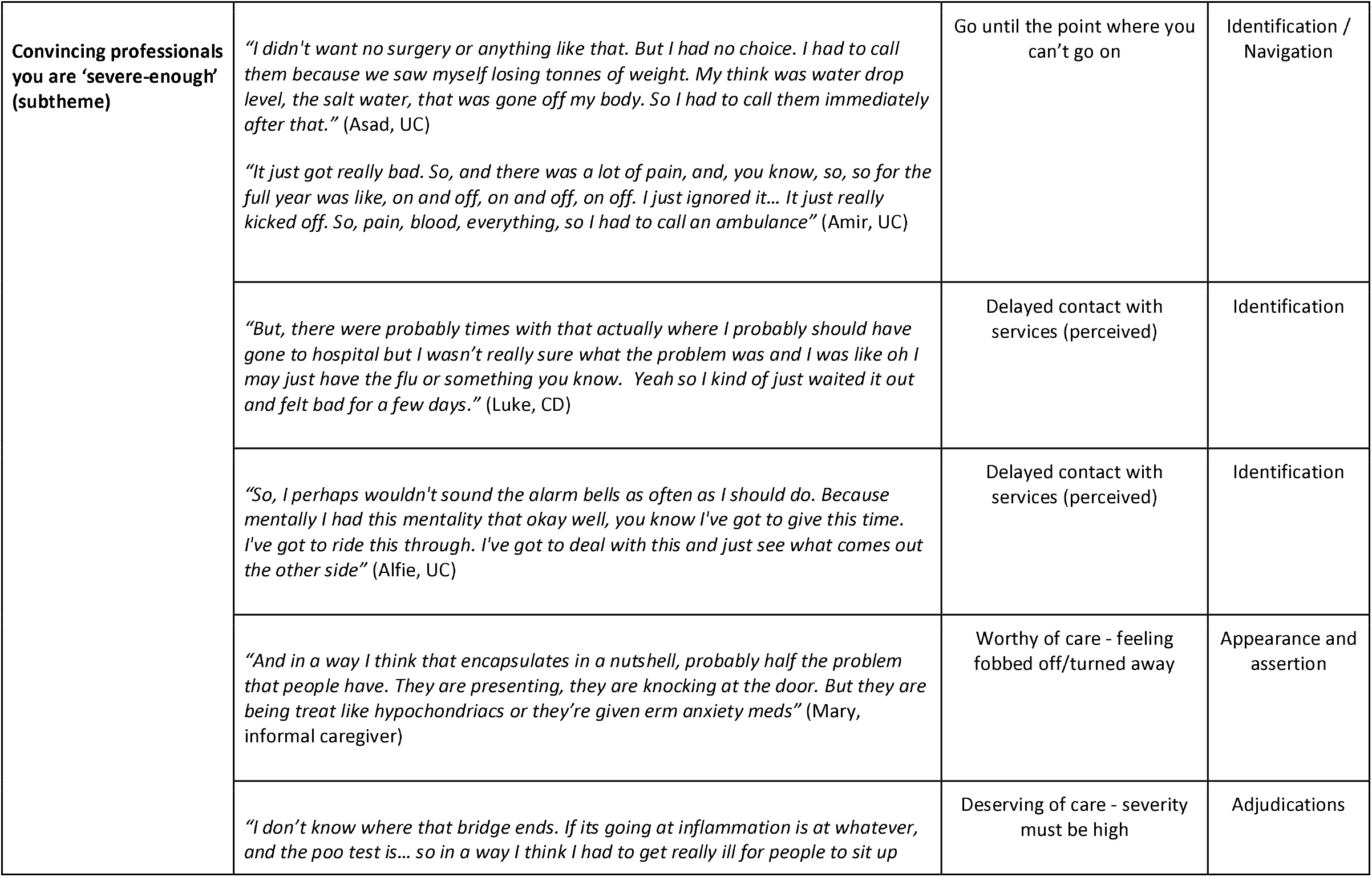
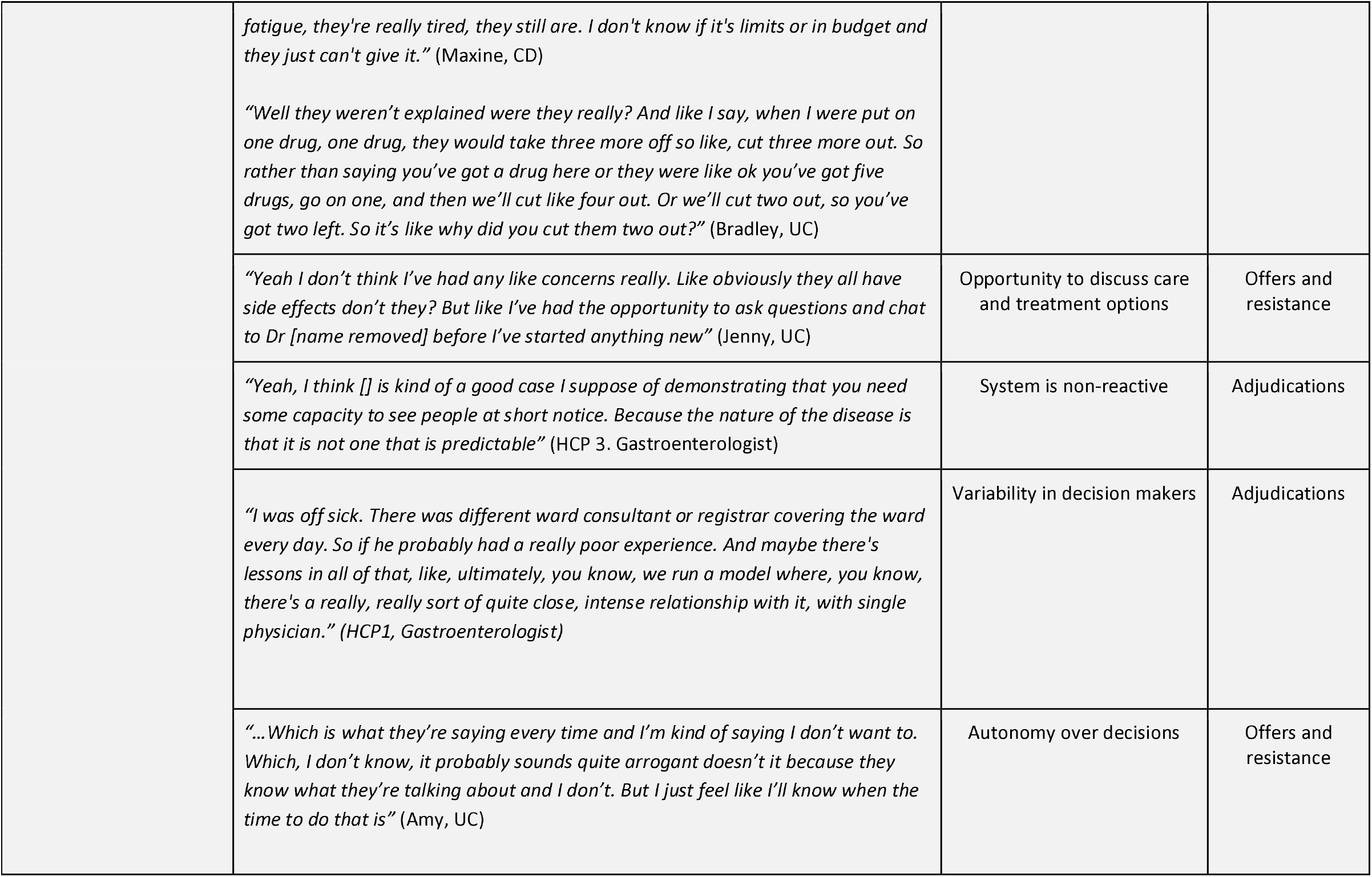
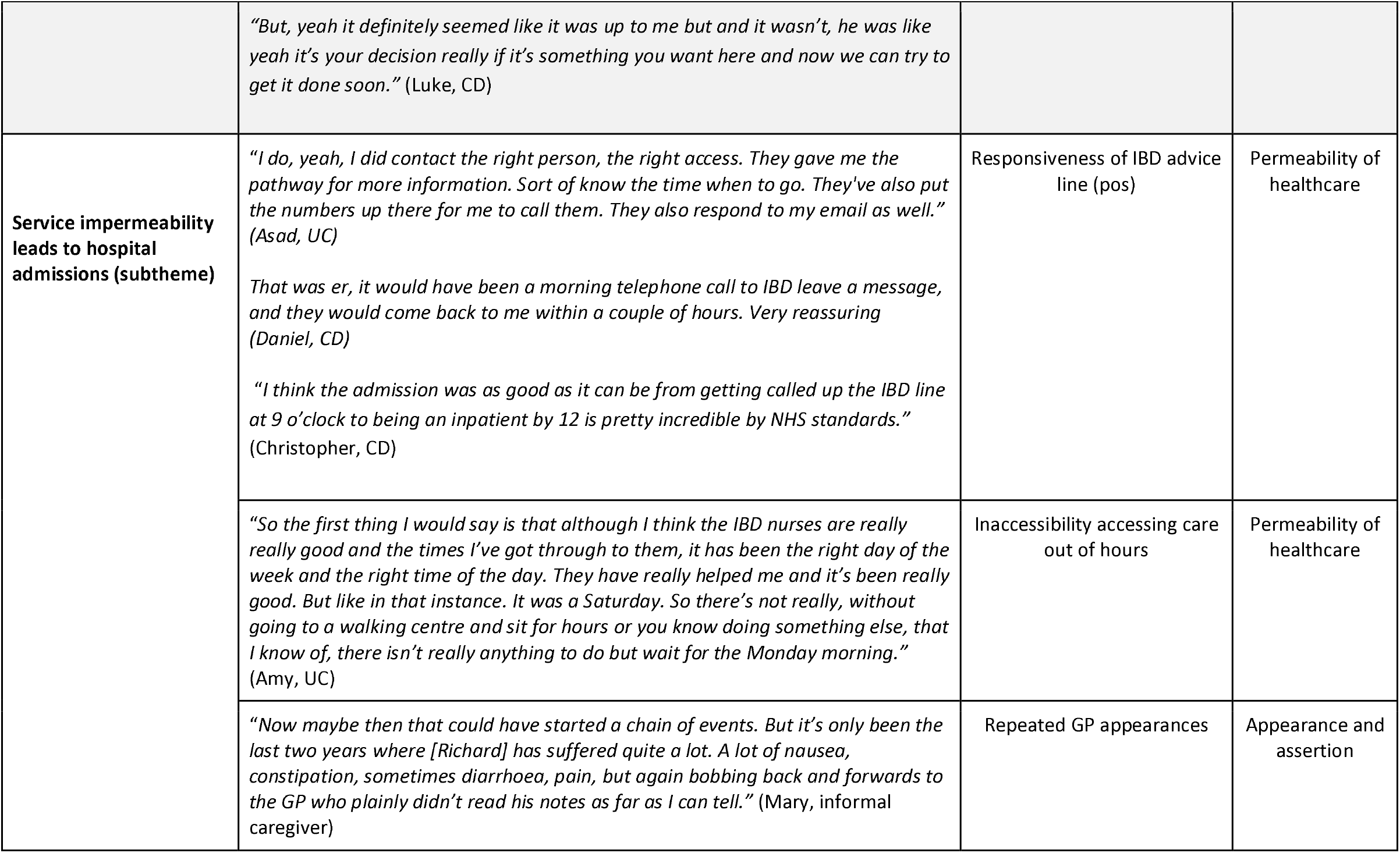
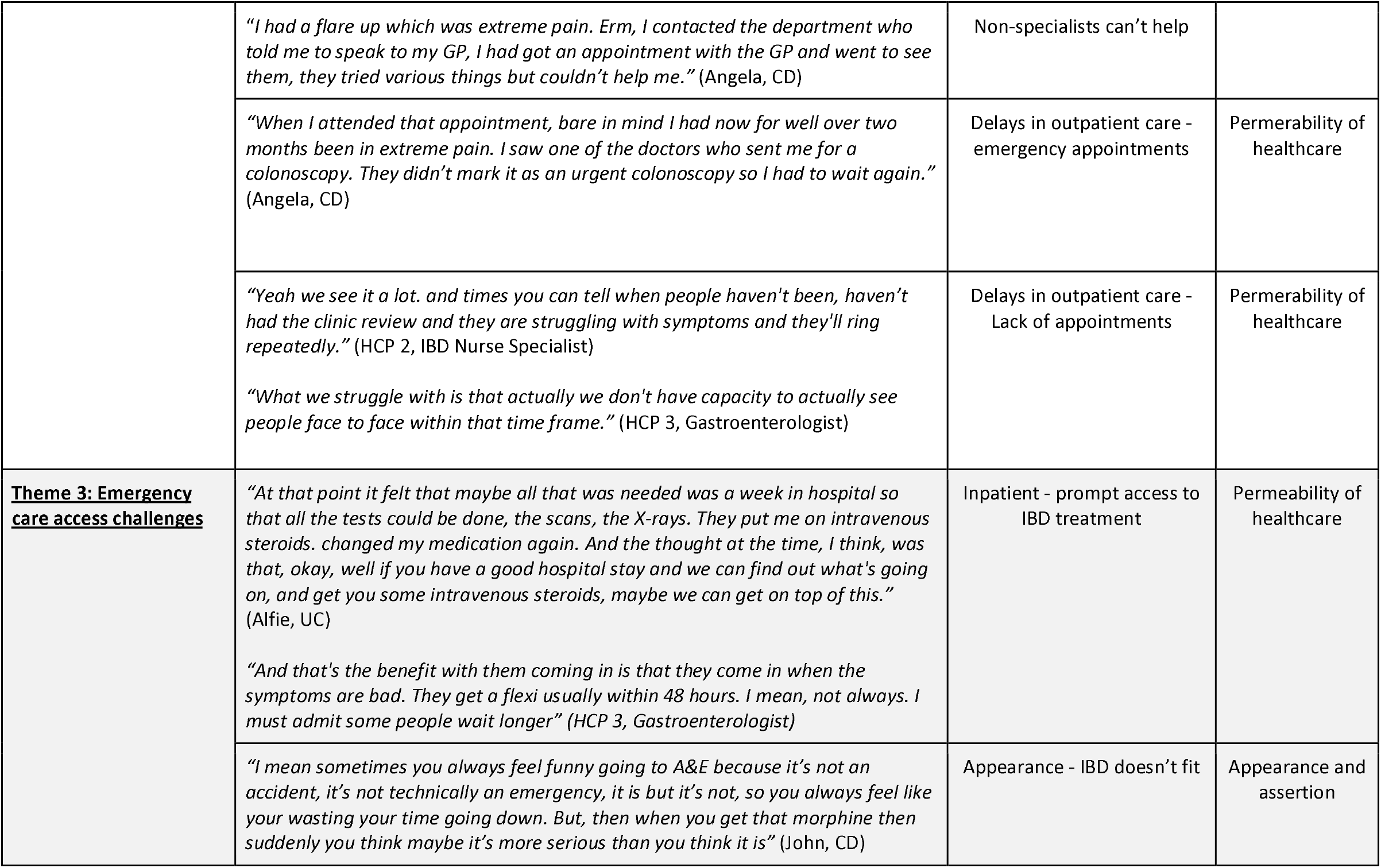
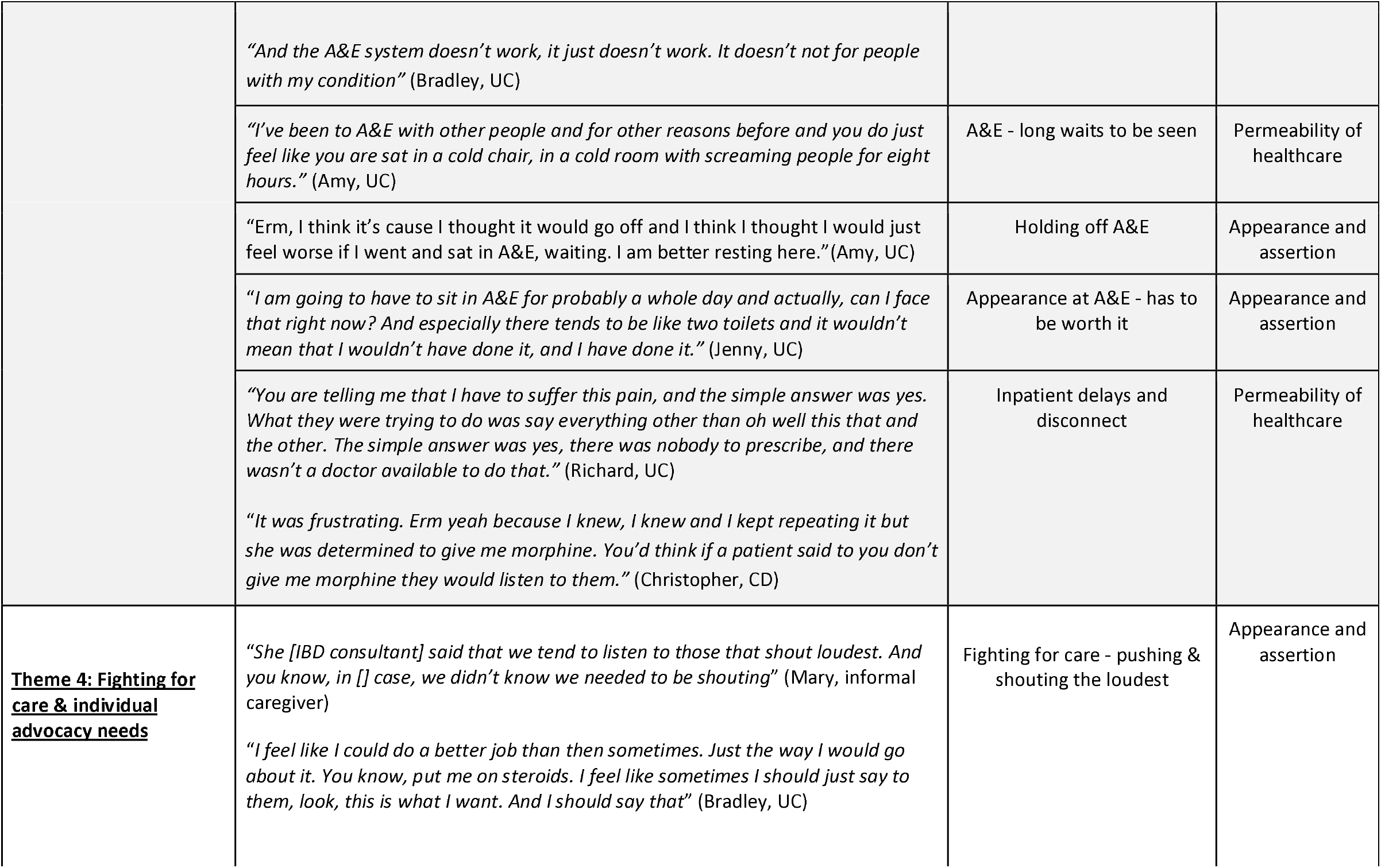
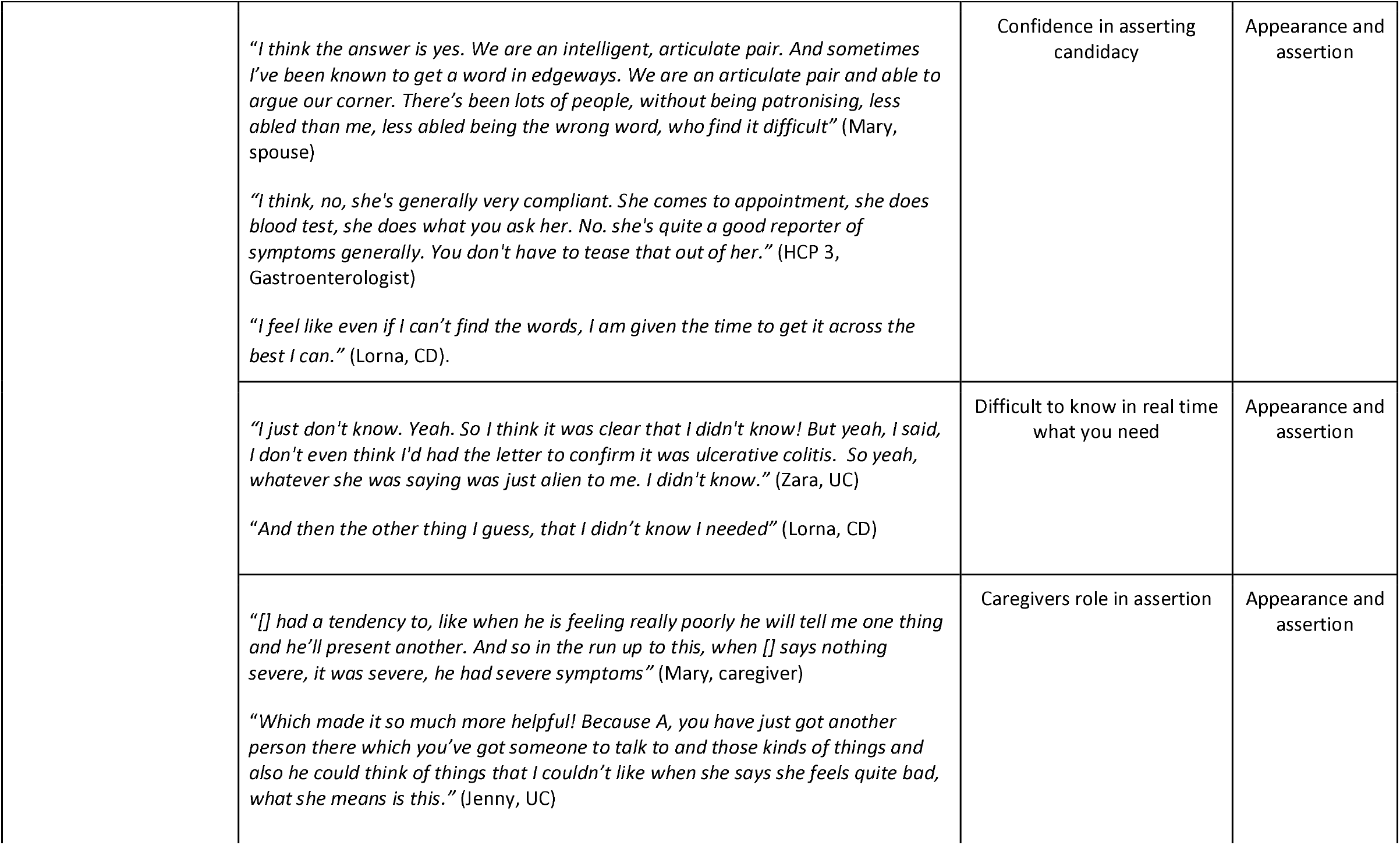
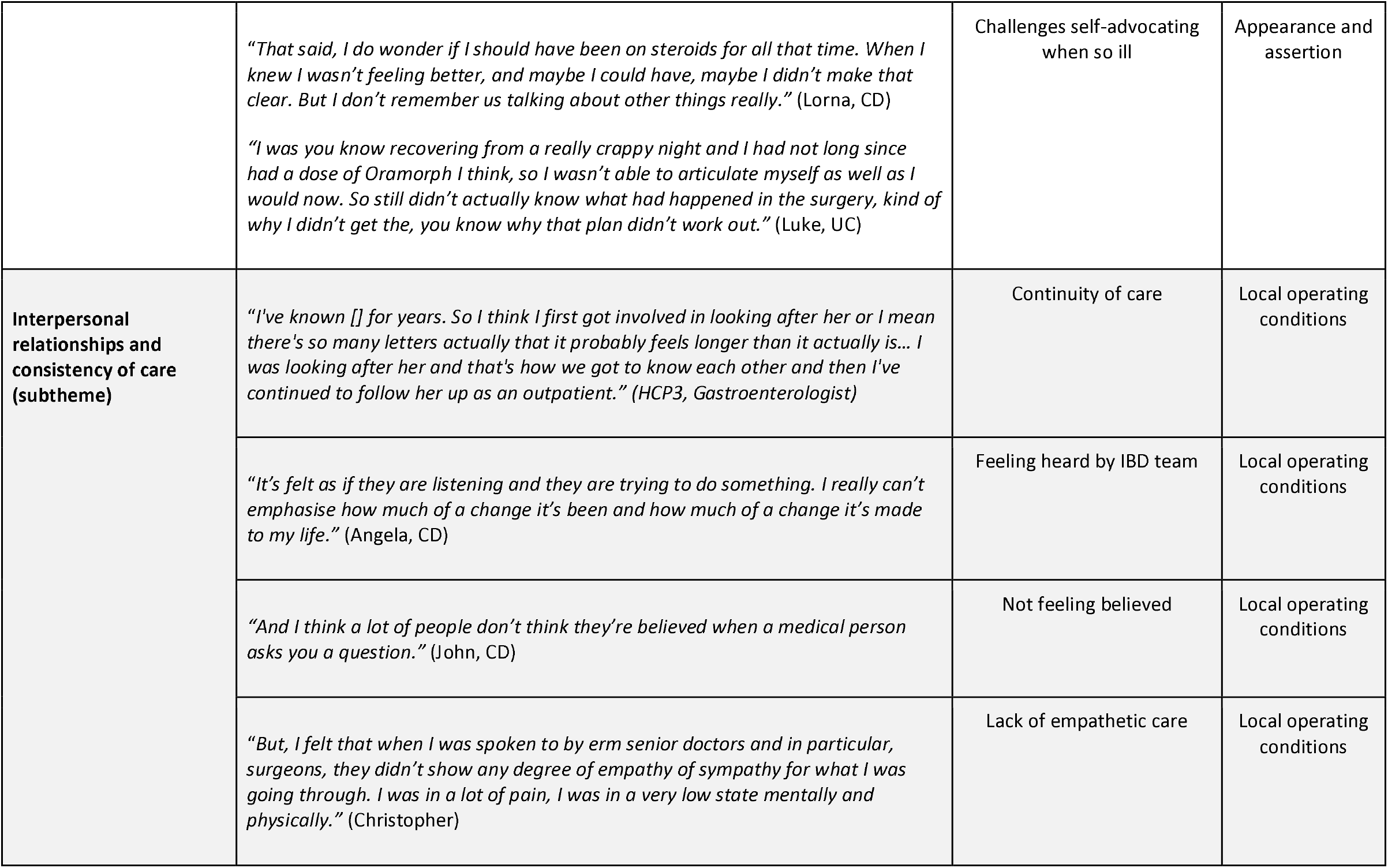
Qualitative themes with quotes.

**Figure 1.**
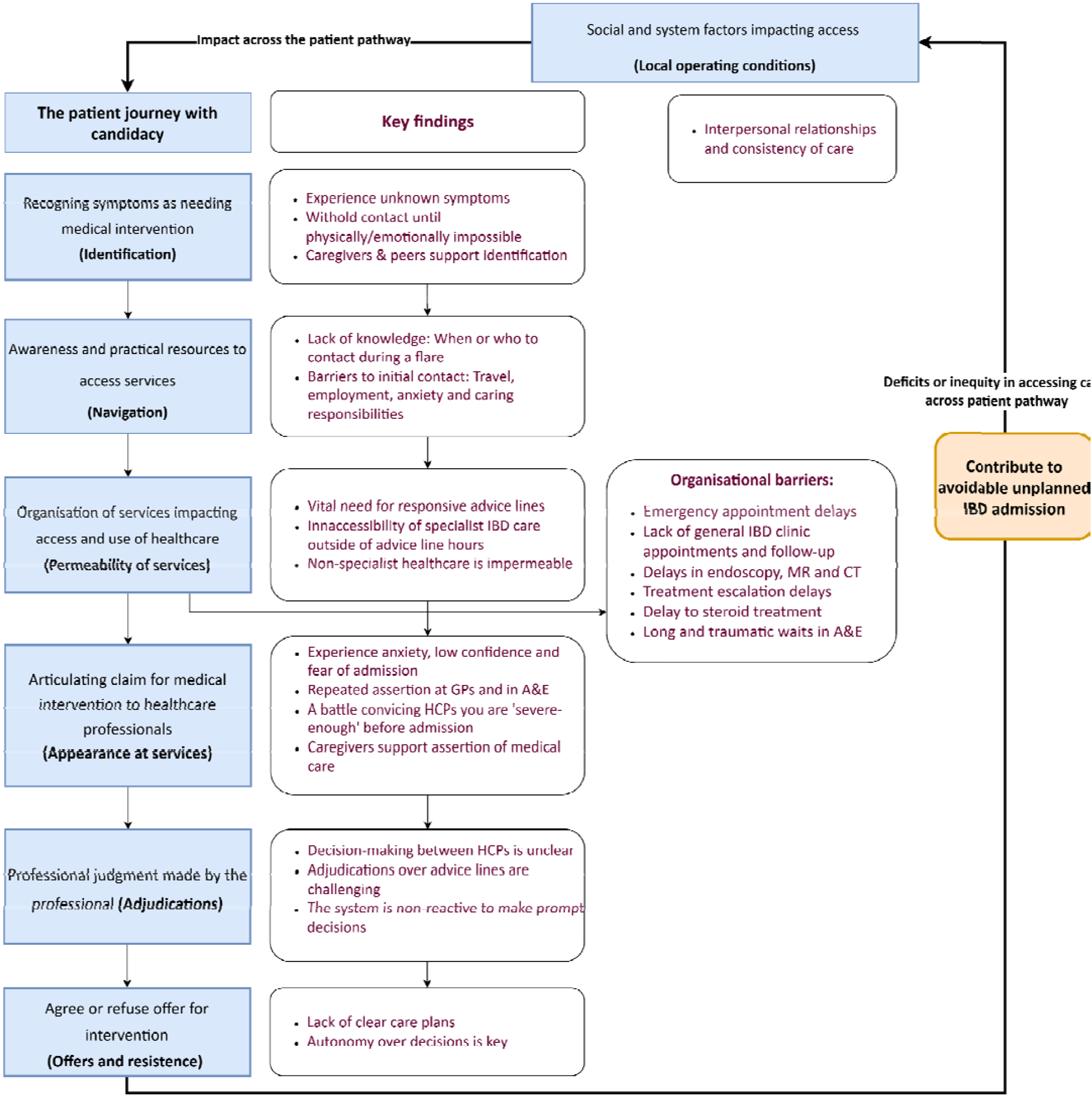
Qualitative findings outlining the patient journey with candidacy in unplanned IBD admissions

#### Theme 1: Difficulties in identifying flares and asserting severity before admission

Participants described the difficult process of recognising a flare and asserting their need for intervention to clinicians before or during the point of hospital admission. Delays in flare recognition and initial contact with services - impacted by flare uncertainty, knowledge of services and confidence in assertion - meant that people with IBD felt they accessed care late and were admitted with more severe symptoms than if they had contacted services earlier.

#### Subtheme: Uncertainty in identifying flares

People with IBD, caregivers, and IBD clinicians described the internal struggle people with IBD experienced in identifying if a flare is happening, in knowing when to seek help for a suspected flare. For people newly diagnosed, identifying a first flare was especially challenging:

> *“I had never had a flare up before, so I didn’t recognise the symptoms before I was admitted, otherwise I wouldn’t have left it as long as I did*.*” (Christopher, CD)*

In this case, Christopher had waited five days before contacting the IBD advice line, feeling as though this was too late. However, the gastroenterologist did not perceive a delay at this point, but earlier on care, highlighting differential perspectives:

> *“So I guess you are maybe going to ask this, but where is the delay here?*… *Before we ever saw him, there was a delay in giving him an ineffective drug treatment for three months before we saw him…The help line wasn’t it, this time I think*.*” (HCP4, Gastroenterologist)*

Differences in patient and clinician perspectives were also highlighted between Luke and his Gastroenterologist. Luke shared that he felt, “*there were probably times with that actually where I probably should have gone to hospital but I wasn’t really sure what the problem was”* (Luke, CD), whilst his Gastroenterologist perceived him at that point when she saw him in clinic as *“pretty well and doing better than he was”* (HCP 5, Gastroenterologist), highlighting discrepant perceptions of disease severity and temporal physical wellbeing between patients and clinicians.

#### Subtheme: Not always knowing when or who to contact

Many people with IBD felt unsure about who to contact for help; highlighting the need for “*people to know a bit more about what the team do and what they cover and a bit more about the condition as well*.*”* (Zara, UC). Clinicians also acknowledged lack of knowledge as a barrier to contacting the service. This was especially true for Richard, who was unaware of his IBD diagnosis for over 10 years, which subsequently resulted in a severe unplanned admission:

> *“No, we didn’t know they [the IBD service] existed. Like I say we didn’t know we had this condition so there was nowhere else to go and that’s why [Richard, spouse] keeps referring to seeing a specialist and paying privately…of course by the time we got to that point he was already very very critical*.*”* (Mary, informal caregiver)

Negative experiences and *“fear of going into hospital”* (Daniel, UC) led many to delay appearance at IBD services or to A&E until they were extremely unwell. A few shared how this decision was based on whether the severity was ‘worth it’ to face the wait:

> *I think I had gone two to three days before I actually went into the hospital to see if I could try and clear it myself. And no, it just doesn’t work. Once it starts, it starts*.*”* (John, UC)

However, most felt more confident to identify future flares and contact the service after their unplanned admission due to improved judgement about their current symptoms based on previous flares. Some gastroenterologists also acknowledged this. In another interview, a participant shared how their risk perception had changed since their admission:

> *“When I called up the IBD nurses for steroids last time I was on, I was going travelling the next day or something. And in my head I was not going travelling without having some support before I went. Because I felt like a flare was coming along and I wasn’t taking that risk. And I was nowhere near as into a flare as when I was before my admission*.*”* (Amy, UC)

Therefore as confidence grows, people with IBD can develop strategies as a safety net to mitigate potentially arising problems associated with the unpredictable symptoms.

#### Subtheme: Practical and emotional barriers impeding timely contact

Travel costs, time off work, loss of earnings and family commitments were also barriers to contacting services earlier before unplanned IBD admissions. This resulted in some participants withholding from contacting the IBD service during their disease flare. For Asad, he felt he *“couldn’t be bothered to go to the doctors just too far away*.*”* (Asad, UC). For Jenny, childcare arrangements remained a concern during the interview:

> *“…But if I’m in hospital for five days then who is going to look after her? So you are trying to like make sure that also works*.*”* (Jenny, UC)

Another participant discussed concerns over loss of earnings:

> *“Because, you know if you’re going into hospital, you’re going in for about a week. And the upheaval it causes at home and everything else. Loss of earnings, I’ve only just started in the job I’m in now, so I lost a week wages out of it. And you try your best not to let that happen because it does impact you*.*”* (John, CD)

Anxiety and fear was perceived to delay calls to the advice line from both people with IBD and clinician perspectives; *“I think anxiety plays a big role for [refers to a patient]. And that’s not uncommon at all”* (HCP 1, Gastroenterologist). Fear of calling the advice line was also discussed in an interview with Asad:

> *“And that’s when I saw blood coming out as well. And I was just afraid to call them, because I really had no energy to dump anything at all. So I just let go. I started doing the same routine, but I lay in my bed until I said to myself, you need to really get up and just go there*.” (Asad, UC)

The importance of caregivers and social support was highlighted. This included peers and family members prompting the person with IBD that they are not well, supporting people with IBD to identify themselves as candidates for healthcare:

> “*I was so knocked out. And one of my very best friends in the world, he pointed out to me, he said, You just made your entire life focus on holding that job down*.*”* (Alfie, UC)

#### Subtheme: Convincing professionals you are ‘severe-enough’

Once at the point of contact with healthcare professionals, many felt reaching a crisis point was necessary before their symptoms reached a severity threshold to qualify for treatment escalation. Some people with IBD also shared they felt delayed in contacting the IBD team and/or in asking for help from family and friends, disclosing they carried on until *“it just got really bad”* (Amir, UC) and it became impossible to carry on:

> *“And then it got to the point where I couldn’t do it anymore. That was the day that I phoned up the IBD nurses and sort of said I can’t keep going like this basically and that was the point that I was first admitted*.*”* (Lorna, CD)

Participants shared a range of challenges in meeting a threshold for treatment escalation from the IBD team, GPs and A&E frontline staff. Some were told to wait up to 8 weeks for treatment to take effect, causing uncertainty. For Bradley, he went back and forth between A&E consultants and on-site GPs when he self-presented to A&E:

> *“And as soon as you say you’ve got an [IBD] consultant they [A&E department} send you straight down the GP. So as soon as that first happened we were thinking amazing, we’re going to be seen. No no no, that’s warning bell. They’re going to see you, they will give you some steroids, maybe, and they will tell you to ring your consultant. And we went back up didn’t we?*… *You had an argument with the GP because you (speaking to Bradley) were bleeding…*” (Lucy, informal caregiver)
>
> ….
>
> *“Yeah because they just want to get rid of you*.*”* (Bradley, UC)

Assessing flare severity over the IBD advice lines was a common challenge discussed by all participant groups. For some people with IBD, the imposed clinical indication of severity misaligned with the physical impact that was experienced. Some clinicians shared the high volume of *“pressure on them [IBD nurses] because they have to make complex judgments over the phone which is not as easy”* (HCP4, Gastroenterologist). An IBD Nurse Specialist echoed these challenges:

> *“I think overall, it’s not just [referring to patient] there’s a lot of patients that you just not quite sure on the phone, but their symptoms you may think okay, they’re not horrendous, and they’re on treatment and we just need to give it time to work but there’s always that element of uncertainty about what if*.*”* (HCP2, IBD Nurse Specialist)

#### Theme 2: Navigating a disjointed healthcare system

This theme covers the practical challenges people with IBD experienced in understanding and navigating healthcare to access the appropriate care they needed before admission. Process and organisational barriers, and outpatient delays prior to the unplanned admissions were perceived as causes to some admissions. These are presented in table 4 (supplementary file F) and included in Figure 1.

**Table 4.** Outpatient barriers perceived as precursors to admissions.

| Barriers perceived as causes of unplanned IBD admissions | Example quote |
| --- | --- |
| Delays in access to emergency outpatient appointments “hot slots” | *“I think we could provide more hot slots, but we proved that the hot slots really help, but we don't really have them. So, I think you know whether [], even though I was off sick, you know, if [] had had a hot slot with someone else, maybe he wouldn't have got admitted” (HCP1, Gastroenterologist)* |
| Lack of available general clinic appointments | *“Because I worry, I lose sleep about the number of patients on my access plan and the fact that I'm asking I'm seeing them in clinic. And saying, I want to see them in six months, and I'm seeing them two and a half years later. and I hate that.” (HCP 5, Gastroenterologist)* |
| Delays in endoscopy, MR, and CT scans | *“There's then a barrier between the endoscopy happening, and people looking at it with the result and saying yeah we can make the decision. And that means getting people getting in front of a doctor but also getting in front of the MDT, because that's our processes to correct that to make that decision” (HCP4, Gastroenterologist)*<br>*“We don't have capacity to get endoscopies within that time frame.” (HCP3, Gastroenterologist)* |
| Treatment escalation delays | *“I think I just saw it on the Monday, and I was like, well, we just need to, like, maybe I was just like, why are we messing around? We'd already said we were going for Upadacitinib. And so, I do remember feeling a bit frustrated, of like, what is going on here? But, and maybe he did too.” (HCP 1, Gastroenterologist)* |
| Delays or lack of capacity for follow-up appointments | *“...but then it's that need to review the response to it and probably if there had been better capacity in my clinic then she probably would have seen me a month or so after starting the Mirikizumab, which would have then meant she didn't need to ring the helpline twice.” (HCP3, Gastroenterologist)* |
| Delay in access to steroid treatment | *“So, erm probably could have been prevented if they put me on steroids and they discussed my treatment sooner with everybody and got it all sorted.” (Bradley, UC)* |

#### Subtheme: Poor communication of care plans

Varied experiences were reported about plans for IBD care, which appeared important for building trust between individuals and their IBD team. Some reported good understanding of their care; *“In my case, there’s not been any confusion”* (John, CD). However, others felt frustrated, isolated and wanted more guidance. Interacting with different professionals (e.g., IBD specialist, GPs, A&E staff) impacted treatment access and care decisions for some people with IBD. This was the case for Zara, who was seeing her GP before being referred to the IBD service, and was then later admitted:

> *“*…*It’s made me reflect on why I wasn’t admitted before when my symptoms were so bad and why the doctor [GP] didn’t send me for a test, I when I first sort of was ringing the doctor about that that year and a half period before…that was the worst it’s been”* (Zara, UC)

Autonomy and the opportunity to discuss treatment options about medication and surgery were examples of positive experiences of healthcare and care plans. However, some people with IBD and caregivers experienced poor communication and felt decision making surrounding treatment was unclear, for some their treatment failed, and then they were admitted. For Asad, when asked on what he wished for more information about when an inpatient, he answered:

> “*I would say the surgery, as there was not really tell me more information about what is actually happening inside. As all the doctors telling me is that your inflammation is high. He told me to stop. Don’t miss, no medication. Those are only the two informations only given me*.*”* (Asad, UC)

All participant groups discussed a lack of transparent decision making in IBD care. Gastroenterologists highlighted this across different areas including in endoscopy and MDT processes. One gastroenterologist highlighted how this makes it difficult to decipher retrospectively if an IBD admission was avoidable or not:

> “*it’s just highlighted really sometimes decision making is not clear. In retrospect, I think maybe the decision could have been made to give him the Anti-TNF on that first colonoscopy, but actually giving him a course of steroids and then giving him the infliximab is not unreasonable. I think if we looked at it in this way with retrospect, we might have all said you should have had the infliximab*.*”* (HCP4, Gastroenterologist)

Lack of clear care plans also impacted people with IBD after hospital discharge. Alfie experienced isolation after their unplanned admission and emergency surgery, which subsequently resulted in an readmission:

> *“*…*But it was coming out of hospital, the follow-up care was not really there. I felt like I had to deal with this and cope on my own. And I did end up having a big flare up with my stoma. I ended up having to be readmitted to my local hospital, under A&E again*.*”* (Alfie, UC)

#### Subtheme: Service impermeability leads to hospital admissions

Whilst many access barriers were discussed, the IBD advice line was consistently praised for its responsiveness, access to advice and reassurance For Asad (UC), he felt that nurses on the IBD advice line *“did try to help me as fast as they can*.*”*, whilst they provided the validation and “*reconfirmation*” (Christopher, UC) many people with IBD needed:

> *“I think when they [IBD nurse] spoke to me and said we will probably need to admit you, let me speak to another nurse and call you back. I was like yes do, please do. Because I knew that I needed something more than a prescription of steroids, I am going to need some care”* (Amy, UC)

However, when the IBD advice line was unavailable out of working hours, people with IBD found this especially challenging and frustrating, who were told to contact A&E which provided its own challenges. Severe pain was a reason for 111 and 999 calls, resulting in A&E attendances for Daniel:

> *“So it tends to be the middle of the night where nothing is working anymore, the pain killers aren’t working, it tends to be like 2-3 o’clock in the morning when I phone 111. Which you know, I can’t get in touch with the team at that time or anything and it’s just to get rid of the pain and then I can think straight*.*” (Daniel, CD)*

When IBD specialists were inaccessible, people with IBD shared that they ‘fell through the gaps’ and that; “*There’s nothing for the middle [meaning there is no other option than A&E and hospital admission outside of service hours] because there’s no care for the middle”* (Maxine, CD). Participants often reported poor experiences of accessing generalist services such as GPs, A&E and calls to 111 when experiencing IBD flare-ups. Some participants described repeated GP appearances to seek help for their IBD:

> *“And so I went to the doctors for that once and they sort of tested and felt around and everything and they advised me to, yeah basically once I got the MRI booked they would investigate whatever it was. But, I went back a couple days later because it was really bad and couldn’t do anything about it*.*”* (Luke, CD)

For clinicians, the lack of service reactivity (e.g. lack of emergency appointments) made the physical opportunity to make timely decisions to prevent admissions a challenge:

> *“it’s almost like you need hot slots, but sort of almost in real time… like, it’s almost even hot slots, like, well, they’re better than nothing, but even, like, a week or two later, at least there’s some access, isn’t there? But actually, in like, it’s right person right time, like every time, and we just don’t deliver a system that’s right person, right time at all*.*”* (HCP 1, Gastroenterologist)

When discussing avoidability of admissions, some expressed frustrations of a lack of service reactivity and issues of impermeability. Many felt that the IBD admissions was the inevitability of the condition:

> “*I don’t think there could have been anything that could have prevented his [Bradley] admissions at any time. Because it doesn’t work like that*.

> *… But, that’s the frustrating thing. The frustrating thing is that it is the only thing that is available. The IBD nurse said, go to A&E. That’s it*.*”* (Lucy, informal caregiver)

#### Theme 3: Emergency care access challenges

This theme covers the barriers experienced in accessing emergency and inpatient care during an IBD flare. Despite A&E and hospital admission being *“the last resort” (John, UC)*, A&E is the default route advised for an IBD flare especially when IBD services are unavailable:

> *“It doesn’t feel like that’s the right place to go but that is kind of the route most of the time*.*”* (Jenny, UC).

All participant groups viewed A&E as a necessary default route, but one where many people with IBD encountered long and traumatic experiences:

> *“…it’s quite scary because you think I needed the care immediately. I needed it sooner than that. And I had to wait seven hours just for someone who’s going into a place where they’ve told you there’s a bed ready for you*.*”* (Maxine, CD)

However clinician and some people with IBD reflected on the positives of hospital admission. For some, this prompted access to specialists and scans which would have taken much longer in outpatient care:

> *“In the inpatient setting, because consultants are involved, we often bypass that, break all the rules and just give the drugs without the screening. So, there are like scenarios where there is faster access” (HCP1, Gastroenterologist)*

Yet, when there was a lack of specialists on duty, this often hindered access to diagnosis or treatment escalation. People also experienced delayed access to appropriate pain relief. An apparent disconnect during inpatient care between specialists and non IBD-specialists experienced by some participants was reflected further during and with one participant, who identified as Autistic:

> *“From that comfort perspective as well and I’m probably extremely sensitive to that bearing in my mind my autism is the fact that I really struggle with changes of environment…But that level of things was quite distressing on top of what I had to deal with”* (Alfie, UC)

#### Theme 4: Fighting for care and individual advocacy needs

This theme represents the high burden of self-advocacy across the participant journey to hospital admission that may be detrimental to vulnerable populations with IBD, impacting on unplanned admissions.

A common challenge highlighted by people with IBD was that they often didn’t know what exactly it was they needed, especially for people new to the condition; *“Most people won’t know what questions to ask, as we didn’t*.*”* (Richard, UC). This was also the case for Fran:

> “*But I’m not sure you always know everything you need every time in terms of support*.*”* (Fran, CD)

Whilst the majority of people with IBD after their admission felt confident to assert their healthcare needs, many still felt they had to push to actually gain ‘approval’ for access. This was expressed across encounters with GPs, during IBD outpatient appointments, in A&E and during inpatient stays. In retrospect for Luke who was experiencing a stricture, he felt he hadn’t asserted how severe his pain was to the IBD team, which resulted several GP attendances before his admission:

> *“I guess I didn’t get across how bad the pain was erm. Oh yeah because it was initially, I called the IBD nursing unit and I said I am having this terrible really bad issue with pain and I don’t really know what it is it’s probably to do with the stricture and, well we can’t really prescribe anything for that but you should go see the GP for that”* (Luke, UC)

For some people with IBD who identified as having a learning disability or as neurodivergent, they shared communication challenges prior to and during their unplanned IBD admission:

> ““*Because [Bradley] is dyslexic, and his communication skills can be, interesting sometimes. Because he’ll go into a meeting with [names consultant] and again he’ll be led by [consultant] rather than this is the context of what’s happened”* (Lucy, caregiver)

Many felt they had to *“fight for care”* (Lucy, caregiver) to have their needs met. However many people with IBD and caregivers highlight how incredibly difficult self-advocacy was during inpatient stays:

> “*I think maybe something to recognise is the fact that if people are in a point where they’re suffering from a debilitating illness, is, what should be straightforward. If you’re living a functional life can be challenging. It strips back your layers of ability to have the cognitive function to put two and two together” (Alfie, UC)*

#### Subtheme: Interpersonal relationships and consistency of care

Across interviews, interpersonal relationships were central across participant experiences of hospital admission. Positive experiences supported people with IBD to; seek appropriate help in a flare, feel heard, understand the IBD service and be willing to engage with it. This was evident for people with IBD who faced existing barriers to access and engage with healthcare:

> *“Them listening to me? It’s because of my of my eating habit, my weight*… *I just wanted the IBD nurse to give me more information…and they just told me to keep taking medication. And from there, I just kept listening to listening to them, and I had to take it every time. I did miss a few medications here and there, but I’ve put myself back up just eat it every time*.” (Asad, UC,)

On the other hand, negative encounters (e.g., being dismissed by a professional, isolation, lack of empathy) left participants without the reassurance of what to do in future IBD flares. For some, negative inpatient experiences increased their hesitancy to engage with IBD services in the future, potentially impacting the trajectory of a future IBD admission and trust:

> *“I think it’s always been at the back of my mind…But there’s always that thought in the back of my mind of what if they just leave me again and erm it does make me a bit hesitant to contact them*.*”* (Angela, CD)

## Discussion

This qualitative study explored the healthcare experiences of people with IBD and some of their informal caregivers and IBD clinicians surrounding unplanned hospital admissions. We identified four central themes illustrating how access across the patient pathway is negotiated, and frequently obstructed, contributing to unplanned IBD admissions.

Our findings revealed the persistent uncertainty experienced by people with IBD during the identification of flares prior to admission, due to lack of knowledge about IBD flare symptoms. The challenges of coping with this uncertainty have been highlighted by previous research exploring inpatient experiences in UC (6). For clinicians, making complex judgments whether to admit or wait over the telephone added an extra layer of uncertainty (26) to this problem, potentially delaying timely access. When comparing perspectives, we found that people with IBD and clinicians often did not align on “timely” contact and “severe” IBD symptoms, meaning it was difficult to decipher whether people with IBD in this study accessed care at the right time or not. Research also demonstrates that patients and professionals are often misaligned in other areas of IBD care including pain discussions (27) and clinic appointments (28). These combined challenges meant that people with IBD would wait to see if symptoms worsened with some delaying contact until they reached a physical or emotional breaking point. These findings highlight the need for improved and standardised information provision about IBD flares and when to contact services in order to minimise and prevent admissions.

The IBD advice lines were consistently praised for promoting prompt access to advice and support and streamlining admissions through a direct route to an inpatient ward for some. Cumulative research supports the positive impact of IBD advice lines on service access and permeability (29,30). However, the impermeability of wider healthcare exacerbated the reliance on emergency services as a default access point. In the UK, many people use emergency departments in order to access healthcare they are unable to elsewhere (31). Participants in this study identified A&E as an access point for quicker tests (e.g., scans, bloods) however as a suboptimal environment for IBD care, describing the process fraught with long waits and a lack of specialist knowledge. Whilst there is an absence of research reporting IBD experiences in A&E, these experiences are echoed in UK charity reports (1,3). Further research is necessary to explore opportunities to improve patient experience in A&E care. The absence of rapid access IBD appointments within the service meant that people with IBD had no alternative but A&E during an acute flare. Therefore, unplanned admissions can result from system-driven outcomes resulting from a lack of reactive, specialist capacity in the community.

Our findings also highlight how certain individuals vulnerable to inequalities may impact how candidacy is asserted. People with IBD shared the high burden of self-advocacy and assertion during flares that may be detrimental to vulnerable populations, including neurodiverse individuals and those with learning disabilities. Research highlights significant health disparities for people with learning disabilities and neurodiversity due to systematic barriers in lack of reasonable care adjustments and poor communication with professionals (32,33). As such, relying on reactive, patient-initiated contact may inadvertently widen inequalities in healthcare access by failing those who lack the resources or health literacy to persistently assert their candidacy (20,34).

The concept of recursivity, defined as “the interdependency between a user’s experiences of health services and their future actions in regard to health and help seeking” *(19,35)* emerged as a critical driver of help-seeking and unplanned admissions. Consistent, empathetic relationships with the IBD team fostered the confidence needed to seek help early for people with IBD. Conversely, negative or dismissive interactions created recursive barriers (19), causing some people with IBD to delay contact until they reached a physical and emotional crisis point. These findings suggest that some delayed presentations to services are recursive responses to prior systemic failures.

### Implications and Recommendations

The study findings carry important implications for improving patient care and reducing unplanned IBD admissions. These recommendations were developed with our PPI contributors. First, these findings highlight the need for IBD services to integrate Patient-Reported Outcome Measures (PROMs) into routine outpatient monitoring (36). Capturing the impact of symptoms may support earlier adjudication of candidacy before patients reach a physical crisis and enable patients to have clearer care plans and greater involvement in their care. Second, there is a need for improved rapid-access pathways that provide a viable alternative to A&E. Establishing “hot slots” can prevent unnecessary hospital visits for diagnostic tests and allow for timely treatment escalation in the community. Third, services should formally embed psychological support within the IBD multidisciplinary team to address the fear, anxiety, and trauma associated with flares and admissions. Addressing these psychological barriers is essential for improving recursive help-seeking behaviours. The emotional and psychological impact of unplanned IBD admissions was commonly discussed across this study however was not the studies primary focus. We aim to publish these findings in a separate article.

### Strengths and Limitations

The multi-perspective design is a notable strength, including dyadic and triadic perspectives from people with IBD, caregivers, and clinicians allowed for a broader understanding of people’s healthcare journey. The application of the Candidacy Framework (14) enabled a nuanced exploration of the individual, interpersonal and organisational drivers of healthcare access surrounding unplanned IBD admissions. The sample of people with IBD was notably diverse across gender, age, IBD subtype, IMD, disability and comorbidity. The inclusion of participants from deprived socioeconomic backgrounds, as well as neurodiverse individuals and those with disabilities, addresses a critical gap in IBD health inequalities research (8). Finally, PPI throughout the analysis phase boosted the quality and rigor of the data interpretation.

Several limitations are acknowledged. This study was conducted within a single UK tertiary IBD service, which may limit the transferability of findings to other healthcare contexts. The study lacked a comparison group of patients without unplanned admissions to evaluate the specific enablers of community-based flare management. There was also a greater weighting of patient perspectives, with a low response rate of IBD clinicians in the service (35%), including nurses, and primary care and emergency department clinicians’ perspectives were missing. Therefore, understanding the perspectives of these professionals is vital for a more comprehensive understanding of how to best manage IBD flares in primary care (e.g., The National Primary Care Diagnostic Pathway for lower gastrointestinal symptoms (37)) and the emergency inpatient setting. Finally, participants were asked to retrospectively recall experiences from up to two years prior, which may be impacted by memory recall challenges.

## Conclusion

In this study exploring perspectives of people with IBD, clinicians and some informal caregivers, participants shared how complex and often overlapping experiences of uncertainty, lack of knowledge, unsatisfactory plans for care and high burden of self-advocacy during unplanned admissions can compound to impact access and care experience. For many, the persistent assertion required to navigate a disjointed system created significant barriers, particularly for vulnerable individuals. Systemic impermeability, specifically the lack of out-of-hours specialist advice and rapid-access appointments frequently forced people with IBD toward A&E care. To potentially reduce avoidable admissions, services must shift toward proactive strategies, including patient education, monitoring tools, specialist rapid-access pathways, and formal psychological support.

## Data Availability

All data produced in the present work are contained in the manuscript

## Supplementary Files

### Supplementary file A

### Supplementary file B: Patient and public involvement across the study

PwIBD contributed to the design and analysis of this research study, all contributors were reimbursed for their time. First, an online meeting was held with four PwIBD recruited via social media and from previous IBD projects. During this session, contributors informed the project design, recruitment strategy and interview guide development. Second, a patient panel, formed as part of the AWARE-IBD project, reviewed the study documentation including the participant information sheet (PIS), drafted interview guide and recruitment materials.

Following the completion of interviews, coding and initial theme generation by RH and CC, two public contributors with lived experience of IBD contributed to interpreting and theming the study results. The lead researcher (RH) met PPI contributors at three points across the project: 1) An introductory meeting, 2) 1-hour remote training session on qualitative data analysis, 2) A 2-hour in-person workshop. Prior to the 2-hour in-person workshop, four initial themes with codes and example quotes were sent to the contributors and contributors attended the session with written notes about this data. During the session, RH talked through the four themes and coloured cards with theme names and quotes were placed on a table. During discussions, recommendations for improving IBD care were co-developed. Then, contributors looked at the data as a whole and cards with themes, codes and quotes were moved around the table to agree on a final thematic structure. Queries about codes were checked by the research by revisiting Nvivo and sharing other examples. The themes were renamed, reorganised to overarching themes and subthemes. For example, emergency care findings were not originally in its own theme but contributors felt that this was important, as it separates from outpatient care. Following the meeting, a final thematic structure was circulated with contributors and themes were agreed.

Embedding PPI across this project, especially during the analysis, boosted reciprocal learning opportunities and improved the quality, rigor and process of reflexivity in the data analysis through triangulating with multiple perspectives (25). The purpose of embedding PPI was to encourage reflexivity to and discussion of the process of conducting the qualitative analysis. PPI contributors sense-checked the data, including the codes and themes against example quotes.

### Supplementary file C: Sampling Frame

## AWARE-IBD Reducing Unplanned Admissions: Patient interviews sampling frame for clinicians

### Document purpose

To support clinicians at the IBD Centre in identifying patients eligible for taking part in an interview about their experiences of healthcare related to unplanned admissions who are not within the existing AWARE-IBD patient cohort.

### Background

To explore experiences prior to and during an IBD unplanned emergency admission. We will recruit dyads or triads of IBD patients and their IBD clinician care team (consultant/IBD nurse). In order to compare experiences, we will also aim to recruit patients who have not experienced an unplanned admission for their IBD.

Participants are being recruited to take part in an interview via the AWARE-IBD cohort and also outside of the AWARE-IBD study. The reason for recruiting outside of the AWARE-IBD cohort is to voice the experiences of those underrepresented in the AWARE-IBD cohort.

### Work package aims

To analyse case studies of patients who have experienced a preventable IBD unplanned admission in Sheffield.

### *Note

The study will also ask permission from the participant to contact the healthcare professional (gastroenterologist, surgeon, nurse, GP) involved in their care, where applicable. This document is focused on recruiting patients to an interview only.

#### Eligibility

Patients will be eligible to take part in this sub-study if:

⍰ Participant of any gender, aged over 16 years old
⍰ Participants must have either a presumptive or confirmed diagnosis of inflammatory bowel disease
⍰ Diagnosis of IBD based on conventional clinical, radiological or histological criteria.
⍰ Have 1 or more ‘risk factor’ identified from the sampling frame
⍰ Have had an experience of an IBD-related emergency admission within the last 2 years.
⍰ Have not experienced an unplanned admission for their IBD despite scoring their IBD control and experience of care to be indicative of poor experience (either from PREM/PROM data or from a verbal reported encounter with a HCP)

##### Sample frame for clinic recruitment

| Patients with unplanned admissions | Patients without unplanned admissions |
| --- | --- |
| Demographic characteristics:<br>- Ethnic minority background<br>- Lower socio-economic background<br>Clinical characteristics:<br>- Comorbidity<br>Care experience indicated during consultations:<br>• Who have reported not feeling engaged, knowing who to contact in their care team, no support at home<br>• Who report good experience of care despite having admissions | Demographic characteristics:<br>- Ethnic minority background<br>- Lower socio-economic background<br>Clinical characteristics:<br>- Comorbidity<br>- Clinical indication of poor IBD control but no admissions<br>Care experience indicated during consultations:<br>• Who have reported not feeling engaged, knowing who to contact in their care team, no support at home |

##### Process of identifying potential eligible participants within clinic

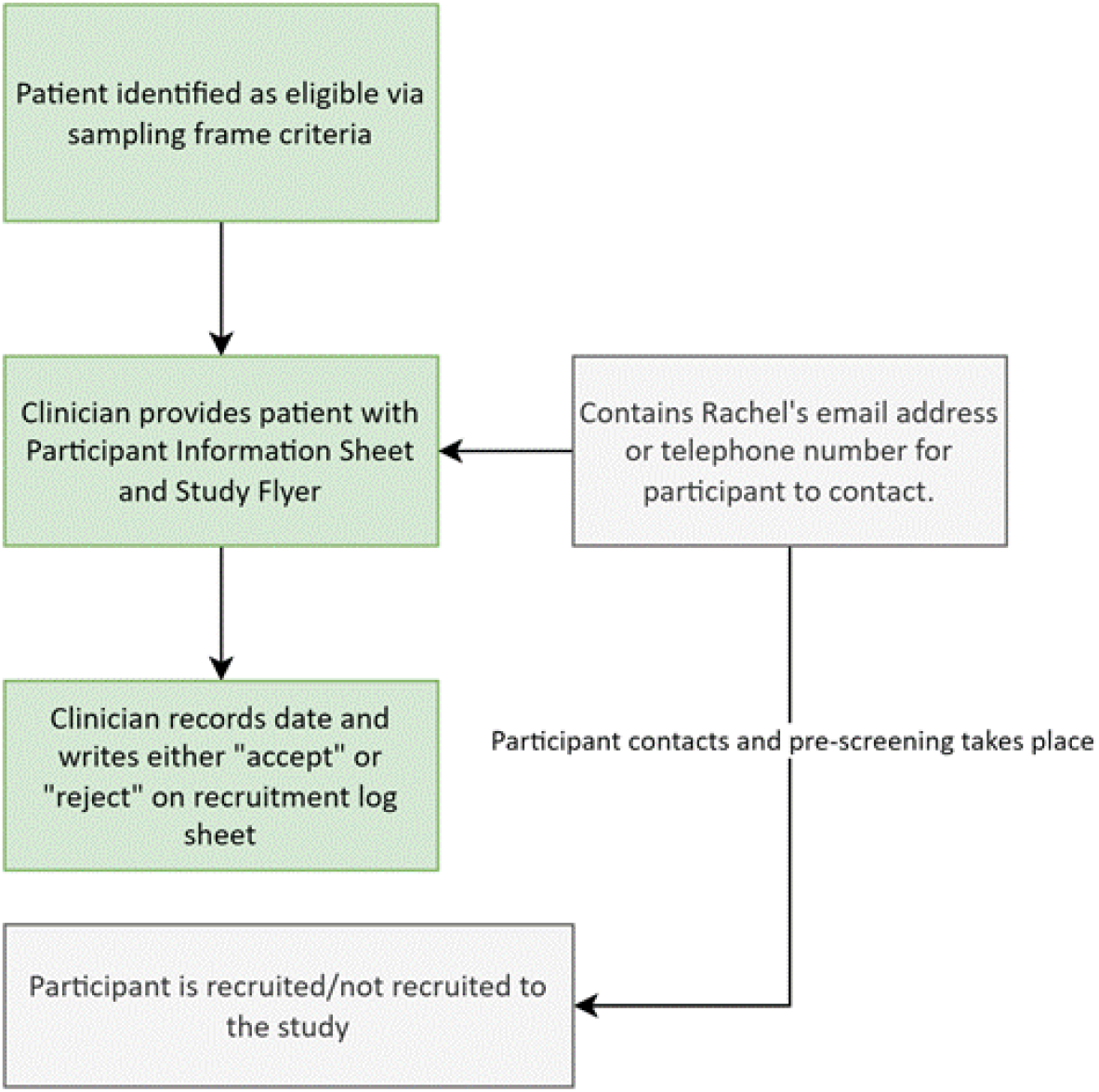

## Supplementary File D: Interview topic guides

### Interview guide: Patients and caregivers

1. If you feel comfortable to do so, can you tell me about what interested you in taking part in this study? <u>Identification and recursivity</u>
2. In your own time, can describe to me your experience of an unplanned emergency admission for your [Crohn’s/Ulcerative Colitis?]
  ⍰ Prompt – when did that happen? (establish timeline)
  ⍰ How long since diagnosis?
  ⍰ How did this make you feel at the time?
3. Can you talk to me about what you were experiencing with your IBD symptoms at the time?
  ⍰ Prompt - how long were you experiencing this for?
4. Can you recall any thoughts or feelings that you had about your symptoms?
  ⍰ Prompts - how severe did you perceive them to be
  ⍰ Did you speak to anyone about these?
  ⍰ Did you seek help from your GP or IBD service straight away?
  ⍰ What information did you need/ Did you have access to any information?
  ⍰ Was there support available?
5. Were the symptoms you experienced similar to a previous flare-up of your disease before?
  ⍰ If yes how were they similar?
  ⍰ If no, what was different about these?
  ⍰ Prompt – any previous vaccination/vaccination history
  ⍰ Prompt – any previous surgeries/ previous biologics/ changes in biologics
6. Can you talk to me about what you decided after experiencing this symptom/pain/change in symptoms/habits?
  ⍰ Prompt/ primary or secondary care
  ⍰ Use of telephone line or other services
  ⍰ Why did you choose to do this?
  <u>Navigation</u>
7. Can you talk to me about what services are you aware of that you can access for support for your IBD during a flare?
  ⍰ Probe - can you talk to me about what your experiences are of using this service?
  ⍰ Would you always contact this service first?
  ⍰ Probe - access/knowledge of the service
  ⍰ Was this service important to you when you experienced you admission?
8. You have talked about how you decided to contact [name HCP/service]. Can you tell me why you chose to contact this provider?
  ⍰ Probe - how easy was it for you to access it/them?
  ⍰ Was this at the appropriate time?
9. Do you feel you had access to the person/service at the right time?
  º Why do you feel this way
  º What could have been different?
  º Why was that important?
10. Did anything get in the way of you seeking this help?
  ⍰ Prompts - what would help you or other patients in seeking help? <u>Adjudications</u>
11. Can you talk to me about what happened when you were first in contact with [named clinician] in relation to your unplanned admission?
  ⍰ Probe: Was a referral needed/ time for referral/ healthcare professional seen at A&E (if appropriate)
  ⍰ Did you see the appropriate clinician straight away?
  <u>Asserting candidacy</u>
12. And when you got into contact with the [relevant person] when you were having the flare for your IBD, did you feel you are able to communicate all your needs to the healthcare professional?
  º Probe: Overall experience
13. Did anything get in the way of you receiving treatment or any other support you needed during this flare?
  º Prompt – any support services? Were they sensitive to your personal needs (cultural/sexual identity)
  º Prompt - signposting? Support offered
  <u>Permeability</u>
14. Did you experience any challenges when trying to access the service?
  ⍰ Probe barriers: Why was this a barrier to you?
  ⍰ What support was needed?
  ⍰ How could this be prevented in the future?
15. [If the participant went straight to A&E] - can you talk to me about what influenced your decision to go straight to A&E?
  ⍰ Probe: perceived urgency, previous experiences
  ⍰ Did try to access any other service/person/support beforehand?
  ⍰ Did you get the appropriate support for your problem?
16. Do you feel services could be easier for people with IBD to navigate?
  ⍰ If yes, how?
  ⍰ If no, can you talk to me about the positives of navigating the service from your experience?
17. How did you feel after your hospital admission? What was the experience of care like for you?
  º If positive, why was it positive?
  º If negative, why was it negative?
  º Have you had a similar experience of being in hospital for your IBD before? If yes, How did this compare?
  º Has how you feel about your care for your IBD changed at all?
18. What could have been done to make your experience of the service better?
  º Probe: Could this have prevented your admission or impact future unplanned admissions?
  º How could the service be improved for patients trying to access it?
  <u>Offers and resistance</u>
19. If you are able to think back before your admissions, are you able to talk to me about any treatment options (including biologics or surgeries) that were communicated to you before your hospital admission?
  ⍰ Probe trust in professional
  ⍰ Probe language/terminology
  ⍰ Probe any barriers to communication
  ⍰ Was understanding of your treatment options checked
  ⍰ What support was available for you?
20. Did you have any concerns about the medication you were prescribed/surgeries you were offered?
  ⍰ Probe concerns about medications/surgeries
  ⍰ Probe any support offered
  ⍰ Do you go ahead/refuse the treatment?
21. Was there any follow up or further treatment offered to you? (e.g. an appointment booked, or a follow up telephone call)
22. Do you think that the service provided you with appropriate support at the right time?
  ⍰ Did you engage with the follow up or treatment? E.g. take medication, maintain contact with the service
  ⍰ What could have been different?
  <u>Operating conditions/local production</u>
23. Reflecting on your hospital admission, do you think that you received the right medical support at the right time?
  ⍰ Has your perspective changed? i.e. from being in hospital to now?
  ⍰ Probe - too early / too late
  ⍰ What could have been different
  ⍰ Why is ‘this’ important to you?
24. Do you feel that your admission could have been prevented or avoided?
  ⍰ Probe – why
  ⍰ what could have changed
  ⍰ what support was needed at the time
25. Is there anything you would change about the service that you used?
  ⍰ Is there anything that could be improved about the service?
26. Would you consider accessing a different health service for your IBD in the future?
  ⍰ If so, which service would you access?
  ⍰ If not, why not?
27. Is there anything else you wanted to discuss in the interview today?

### Interview guide: Clinicians

1. Can you tell me about what your professional role is, and how long you have been working in this role?
2. What interested you in taking part in this study today? <u>Present case study</u>
  · Researcher will spend some time explaining the case study from a patient interview that outlines the timeline of events from their perspective of an unplanned emergency admission for IBD
  · Allow time for participant to ask questions about the case – get patient notes <u>Identification and adjudications</u>
3. Can you talk to me about what happened when you first became aware that this patient needed some support or help for their [Crohn’s/Ulcerative Colitis]?
4. Can you talk me through what decisions you made when you encountered the patient?
  º Probe - challenges in decision making
  º What made you decide ‘this’?
5. Are you able to talk to me about any similar cases to this?
  º If yes, did a similar case before impact your decision?
6. Do you feel the patient was seen at the right time?
  º Probe – to early/too late – why?
  º What could have been different
  <u>Navigation</u>
7. If a patient is experiencing a flare at your service, who would they contact first?
  º Probe – the process
  º Process – is this easy to navigate?
  º Process – could it be improved / challenges
8. Do you feel that the patient’s knowledge of how to access the service was important in this case?
  º Probe – Anything else important here?
  <u>Permeability</u>
9. Were you able to offer support to the patient when they accessed the service for their problem?
10. Do you feel that the patient could have received any support earlier for their problem?
  º If yes – could this have changed the outcome? (admission)
11. Did you experience any challenges when supporting the patient with their problem?
  º Probe - service barriers
12. Do you feel services could be easier for people with IBD to navigate?
  º If yes, how?
  º If no, can you talk to me about the positives of navigating the service from your experience?
  º If yes – is this a problem locally/nationally?
  <u>Offers and resistance</u>
13. Had you previously discussed any treatment options (biologics/surgeries) with the patient?
  º Probe changes in previous treatments
  º Vaccination history
  º Previous surgeries
14. Did the patient discuss any concerns or resistance towards this treatment option?
15. Did the patient need any additional support to make a decision about their treatment plan? <u>Adjudications</u>
16. Who are the important decision makers when it comes to a patient being admitted as an unplanned emergency for IBD?
17. What are the challenges you experience when making a decision about if a patient should be admitted or not?
18. Did you experience any challenges in judgment or decision making during this case?
19. Do you feel that different decision making or judgements earlier on in could have prevented this admission?
20. What are the barriers for clinicians when making decisions around IBD care and management?
  º Follow up – What could reduce or minimise this barrier?
  <u>Operating conditions/local production</u>
21. Reflecting on this patient’s case of a hospital admission, do you think they received the right medical support at the right time?
  ⍰ Probe - too early / too late
  ⍰ What could have been different
  ⍰ Why is ‘this’ important?
22. Do you feel that this admission could have been prevented or avoided?
  ⍰ Probe – why
  ⍰ what could have changed
  ⍰ what support was needed at the time
    º If no – why not
    º If yes – what factors do you feel are important? How?
    º How could services be changed to intervene earlier?
23. Is there anything you would change about the IBD service to support earlier intervention in IBD?
  ⍰ Is there anything that could be improved about the service?
24. Is there anything else you wanted to discuss in the interview today?

### Supplementary file E

### Supplementary file F

